# Mendelian randomisation and colocalisation reveal pleiotropic effects of *CD40/SLC12A5* locus on CD40 protein, depression, and immune disease

**DOI:** 10.1101/2025.08.21.25333931

**Authors:** Rachel Laattoe, Elina Hypponen, David Stacey, Sarah Cohen-Woods

**Author notes:** Denotes equally contributing senior authors (David Stacey, Sarah Cohen-Woods). Corresponding authors: Rachel Laattoe and Sarah Cohen-Woods, Postal address: Flinders University, College of Education, Psychology, and Social Work, Social Sciences North, Sturt Road, Bedford Park, SA 5042, Australia.

## Abstract

Inflammatory pathways are implicated in depression, but the specific immune proteins and causal variants involved remain unclear. This study investigated potential causal relationships between 91 immune-related plasma proteins and depression using generalized summary Mendelian randomisation. We identified a robust association between CD40 protein levels and depression (OR: 0.95, 95% CI: 0.94 - 0.97, *p* = 1.71 × 10⁻¹¹), primarily driven by *cis-*acting variants. However, pairwise statistical colocalisation analyses of the *CD40* locus indicated that CD40 protein and depression had distinct – though not independent – lead variants, suggesting the Mendelian randomisation signal was confounded by linkage disequilibrium. Analyses using expression quantitative trait locus data from the Genotype-Tissue Expression project prioritised *SLC12A5*, not *CD40*, as the most likely effector gene for depression risk at the locus. *SLC12A5* encodes a potassium chloride co-transporter preferentially expressed in brain tissue, consistent with a role in depression. A phenome-wide association study showed the CD40 protein lead variant was primarily associated with inflammatory disorders, while the depression lead variant was more strongly linked to psychiatric conditions. Our results emphasise the importance of combining Mendelian randomisation with colocalisation analyses to disentangle pleiotropic effects at loci with complex genetic architecture, such as *CD40/SLC12A5*. While plasma CD40 protein levels are unlikely to play a causal role in depression, *SLC12A5*-mediated effects may contribute to its pathophysiology. These findings highlight the need for further functional and multi-omic studies to clarify immune-brain interactions and identify therapeutic targets for depression.

## Introduction

Depression is recognized as one of the leading causes of disease burden, affecting over 280 million people worldwide [1]. Despite this, the genetic and biological mechanisms that contribute to the disorder are still not fully understood. Several causal mechanisms have been proposed [2], including inflammation which is shown to be elevated in a subset of individuals [3–6]. Increased concentrations of circulating pro-inflammatory biomarkers including C-reactive protein (CRP), interleukin 6 (IL-6), and tumour necrosis factor alpha (TNF-α), have been reported in individuals with depression and depressive symptoms, in both cross-sectional and longitudinal studies [7–9]. It remains uncertain whether these observational associations reflect causal effects of inflammation, or residual confounding from unmeasured factors such as comorbid inflammatory conditions or environmental factors that trigger inflammation. Genetic approaches provide a potential means to address these uncertainties. The latest genome-wide association study (GWAS) identified 243 risk loci associated with depression [10], several of which are linked to immune-related processes and pathways [11–14]. Gene-based analyses highlighted genes implicated in neuroinflammation and immune regulation [15–18] including *LRFN5*, *GRM5,* and *CD40*. While suggestive of a causal role for immune-related genes, most depression-associated variants reside in non-coding regions of the genome, which makes it challenging to pinpoint the precise causal genes at these loci [19,20]. Genetic methods that incorporate functional information (e.g., gene expression, protein abundance) may help to determine whether immune or other genes are driving depression risk at these loci.

Recently Mendelian randomisation (MR) methods have been used to leverage the increasing availability of functional data [e.g., 21–24]. MR uses natural genetic variants robustly associated with an exposure of interest as instrumental variables to estimate the causal effect of that exposure on an outcome of interest. In this way MR is less susceptible to confounding that may otherwise compromise findings from observational studies. Previous MR studies investigating the relationship between inflammatory protein markers and depression have reported mixed results. For example, a multi-variable MR analysis of immunological proteins found genetically predicted IL-6 associated with an increased risk of major depressive disorder [25]. Other studies, however, reported no evidence consistent with causal relationships between IL-6 signalling, plasma CRP levels, and depression or depressive symptoms [26–28]. Highly multiplexed proteomic platforms (e.g., Olink®, SomaScan®) have recently emerged, prompting several well-powered protein quantitative trait locus (pQTL) studies and a wealth of publicly available pQTL data [29–35]. In combination with high-throughput MR methods, such as generalised summary-based MR (GSMR) [36], these pQTL datasets may help to disentangle the role of inflammatory proteins in depression.

To gain further insight into the causative role of immune proteins in depression we used the largest available Olink inflammation-targeted pQTL [30] and depression GWAS summary data [10] to conduct a two-sample MR analysis using GSMR. We investigated causal associations between the abundance of 91 inflammation-related plasma proteins and risk for depression in > 1 million individuals (294,322 with depression) and conducted comprehensive sensitivity and validation analyses. We identified one signal implicating cluster of differentiation 40 (*CD40*) as a potential causal immune-related protein for depression. However, colocalisation analysis at this locus revealed distinct causal variants for the two traits, with a neighbouring gene, solute carrier family 12 member 5 (*SLC12A5*), also known as potassium-chloride cotransporter 2 (*KCC2*), emerging as the likely effector gene for depression. Follow-up analyses supported this interpretation, suggesting that the *CD40*/*SLC12A5* locus may modulate risk for both depression and immune diseases through distinct mechanisms.

## Methods and materials

### Study design

The study pipeline and methods are summarized in Figure 1 and follow guidelines provided in the Strengthening the Reporting of Observational Studies in Epidemiology Using Mendelian Randomization (STROBE-MR) Statement [37]. Briefly we used a two-sample GSMR design to test for potential causal associations between inflammation-related blood plasma proteins and depression risk. We then used additional MR methods to test the robustness of main analysis findings. Effects from the discovery GSMR were followed up for replication in an independent pQTL dataset. Colocalisation analyses were used to identify any confounding effects from linkage disequilibrium (LD), and a phenome-wide association study (PheWAS) was used to identify other phenotypes associated with the sentinel SNPs.

**Figure 1.**
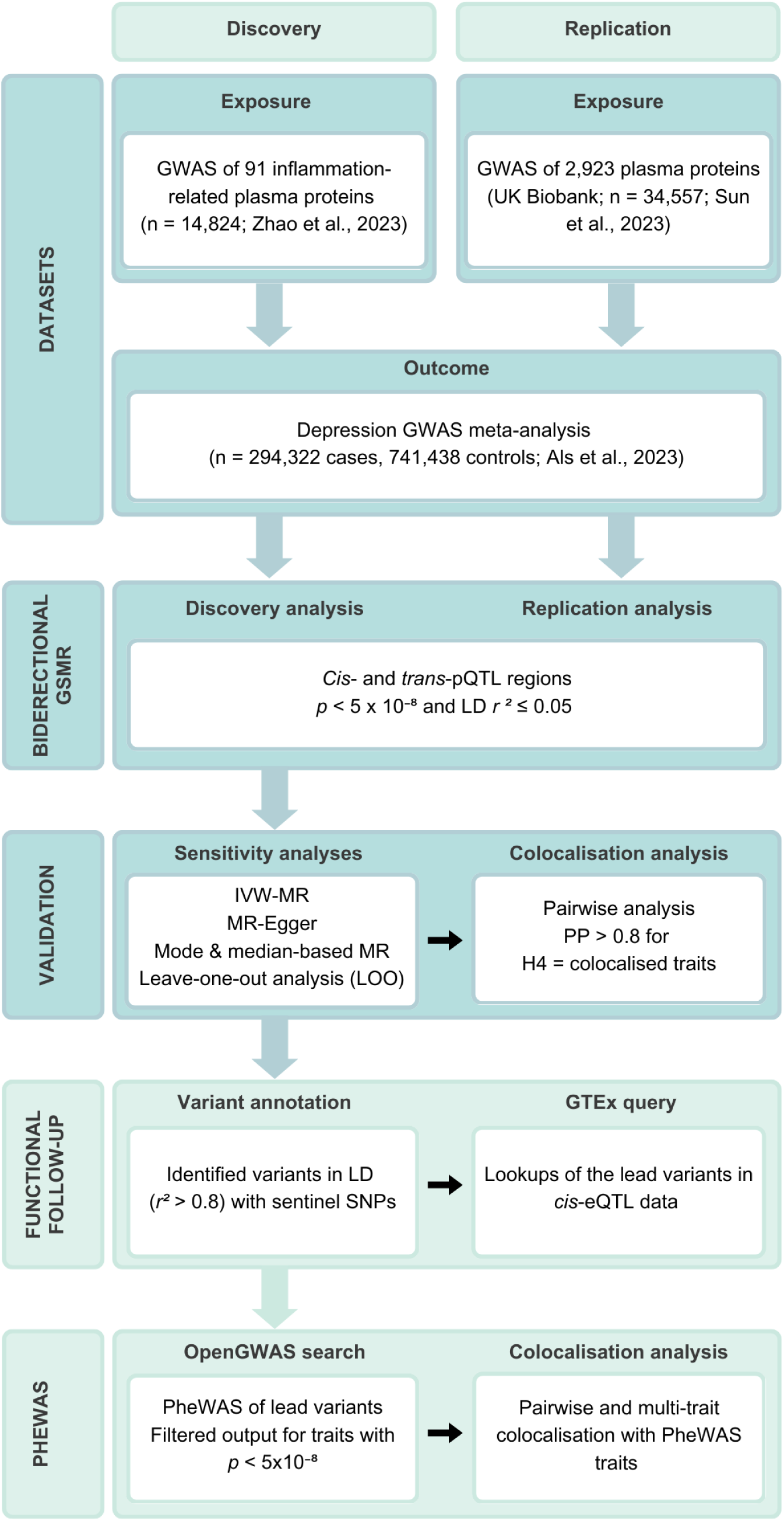
Overview of study design integrating genetic, causal inference, and functional analyses. Schematic illustrating key analytical steps: (i) selection of genetic instruments from immune-related pQTL summary statistics; (ii) Mendelian randomisation to evaluate causal effects on depression; (iii) sensitivity and colocalisation analyses to validate findings and assess shared genetic architecture; (iv) functional annotation of candidate variants; and (v) phenome-wide association study (PheWAS) to identify downstream trait associations and effector genes at the *CD40/SLC12A5* locus.

### GWAS summary statistics

#### Inflammation pQTLs

For the discovery GSMR exposure instrument we used publicly available summary statistics from a pQTL meta-analysis (n = 14,824) of 91 inflammatory plasma proteins measured through the Olink Target Inflammation panel [30]. This included a total of 180 significant locus-protein pairs, comprising 59 *cis-*pQTL (+/-1 Mb of the protein-encoding gene) and 121 *trans-*pQTL regions. Replication analyses were conducted using the matching pQTL data from the UK Biobank (UKB; n = 34,557) [29]. Both the discovery and replication pQTL data were derived from cohorts of European ancestry.

#### Depression

We extracted summary statistics from the latest, publicly available depression GWAS meta-analysis [10] to derive instrument-outcome association data for both the discovery and replication analysis. The dataset, downloaded from the iPSYCH website, included the expanded iPSYCH2015 and FinnGen cohorts, in addition to the Psychiatric Genomics Consortium (PGC), UKB and Million Veteran Program (MVP) cohorts (excluding 23&Me). The GWAS summary statistics were derived from 294,322 individuals with ‘broadly’ or ‘narrowly’ defined depression and 741,438 controls, all with European ancestry.

### Two-sample MR analysis

#### Discovery GSMR

We used GSMR (v 1.94.1) [36] analyses to explore genetic evidence for a causal association between the inflammatory proteins and depression. GSMR, which is part of the Genome-wide Complex Trait Analysis software package [38], builds on existing summary data-based MR (SMR) methods by accounting for the sampling variance for each SNP across both the estimated effect on exposure (bzx) and outcome (bzy), and LD between SNPs via reference sample. Here we used the 1000 Genomes Project (EUR) reference dataset [39] for LD estimation across all analyses. For quality control we applied the default threshold of 0.2 to filter out SNPs with large differences in allele frequency between the GWAS summary data and reference sample. All approximately independent (*r*^2^ < 0.05) variants associated with a given protein (*p* < 5x10^-8^) were selected as instruments using LD clumping, with the minimum number of instruments required for analysis set to 1. Another feature of GSMR is the HEIDI-outlier method, which detects and removes pleiotropic SNPs prior to analysis [36]. As pleiotropy can potentially inflate MR estimates we opted for the modest *P-*value threshold of 0.01, to both conserve power to detect heterogeneity and reduce the likelihood of instruments being removed due to chance [36].

We first conducted discovery GSMR analyses across all 91 inflammation-related proteins, including both *cis-* and *trans-*pQTLs as instruments, followed by separate analyses focusing on *cis-*regions (cognate gene coordinates +/-1 Mb) only to minimize the potential for confounding by horizontal pleiotropy [30, 40]. We defined significant MR associations using a false discovery rate (FDR) of 0.05, calculated by the Benjamini–Hochberg method, to correct for the total number of proteins tested. Following the discovery phase, we conducted a reverse GSMR analysis (--gsmr-direction 1) to account for a potential bidirectional relationship between depression and the significant protein (CD40; *p* < 0.01) identified in the *cis-*only analysis.

#### Sensitivity analyses

To evaluate the robustness of our primary GSMR findings, we performed additional sensitivity analyses using the MendelianRandomization (version 0.3.0) package [41] in R version 4.3.1 (https://www.r-project.org). We implemented inverse-variance weighted Mendelian Randomization (IVW-MR), MR-Egger, median-based, and mode-based methods. These sensitivity analyses focused on the single significant inflammatory protein identified through the GSMR *cis*-only analyses (CD40), which was instrumented by 14 SNPs. Depression GWAS summary statistics served as the outcome. Prior to analysis, we harmonised the SNP instruments across exposure and outcome datasets to ensure consistency in allele frequencies and effect directions. We included an LD correlation matrix generated via the ‘ieugwasr’ R package (version 1.0.1, https://github.com/MRCIEU/ieugwasr) using the 1000 Genomes EUR reference panel to account for residual correlation among SNPs. Leave-one-out (LOO) analyses were used to assess whether any individual SNP disproportionately influenced the observed effect estimates.

#### Replication study

To validate our discovery findings, we utilised summary statistics from the UKB Pharma Proteomics Project discovery cohort [29], which included pQTL mapping of 2,923 plasma proteins in 34,557 individuals using the Olink Explore 3072 platform. We performed combined cis- and trans-, as well as cis-only, GSMR analyses using UKB CD40 pQTL data as the exposure and depression [10] as the outcome. We applied the same settings (including instrument selection) outlined above for the discovery GSMR.

### Statistical colocalisation

We conducted pairwise colocalisation analyses to identify any putative causal signals emerging from MR that may have been confounded by LD. This was implemented through the ‘coloc.abf’ function featured in the R ‘Coloc’ package (version 5.2.3) [42]. For these analyses we used the default prior probabilities, p1 = 1 x 10^-4^, p2 = 1 x 10^-4^, p12 = 1 x 10^-5^, to test each of the following four hypotheses: H0 = there is no causal protein-disease variant; H1 = the causal variant is for the protein only; H2 = the causal variant is for depression only; H3 = there are two distinct causal variants, one for exposure and one for outcome; H4 = both exposure and outcome share the same causal variant. We used the *cis-*pQTL summary statistics for the *CD40* locus (chr20:43747447 – 45758258, hg19) and corresponding genomic region from depression summary data. Traits were considered colocalised if they had a posterior probability (PP) > 0.8 for H4 under the assumption of a single causal variant.

### Depression and CD40 lead variant annotation

We annotated the CD40 protein and depression lead variants along with any other variants in high LD (*r*^2^ > 0.8) at the *CD40* locus using the Ensembl Variant Effect Predictor (VEP) [43]. This allowed us to identify protein altering SNPs in LD with the sentinel variants. We also conducted lookups of the lead variants in *cis-*eQTL data from the Genotype-Tissue Expression (GTEx; version 8) project using the GTEx portal. We extracted all eQTL associations (q < 0.05) involving protein-coding genes.

### Genotype-Tissue Expression (GTEx) eQTL analysis

We conducted pairwise and multi-trait colocalisation analyses to assess shared genetic aetiology between depression/CD40 protein levels and *cis-*eQTL signals from the GTEx project using the R packages Coloc [42] (as described above) and HyPrColoc (version 1.0) [44], respectively. We extracted the relevant GTEx *cis-*eQTL summary data for the *CD40* locus (chr20:43747447 – 45758258, hg19) from the eQTL catalogue [45, 46] – we used the datasets imported directly from GTEx (v8), not the uniformly processed and reanalysed datasets. For both Coloc and HyPrColoc, we considered a PP > 0.8 (PP_H4_ for Coloc) as robust evidence of colocalisation.

### Phenome-wide association study (PheWAS)

We conducted a PheWAS of the two sentinel variants for *CD40* and depression (rs1883832 and rs9074, respectively) using the ‘phewas’ function from the ‘ieugwasr’ R package, which interfaces with the PheWAS database available through the Integrative Epidemiology Unit (IEU) OpenGWAS project (http://gwas-api.mrcieu.ac.uk). We filtered the PheWAS output for neuropsychiatric and immune-related traits/datasets with *p* < 5 x 10^-8^ and selected the lowest p-value in the case of repeated traits (**Supplementary Table 2**).

To examine potential shared genetic variants between traits, we conducted pairwise colocalisation analyses using the ‘coloc.abf’ function on filtered PheWAS traits for CD40 and depression. For each trait, we extracted GWAS summary data restricted to the *cis-CD40* region (chr20: 43747447 - 45758258) from the OpenGWAS resource via the ‘phewas’ function in the ieugwasr R package. Colocalisation was defined as a posterior probability greater than 0.8 for H4. For visual comparison, traits associated with one sentinel variant but not reaching genome-wide significance for the other were also included. Additionally, we performed multi-trait colocalisation for both *CD40* and depression using the HyPrColoc R package [42].

## Results

### CD40 plasma levels are associated with depression

We conducted a two-stage (discovery and replication), bidirectional, two-sample GSMR analysis to investigate causal effects between 91 inflammatory plasma proteins and depression risk. In the discovery stage, *cis*- and *trans*-pQTL variants were used to instrument protein exposures, and outcome summary statistics were derived from an independent depression GWAS dataset (see Methods). After Bonferroni correction for 91 tests (*p*_GSMR_ threshold = 5.45 x 10^-4^), a genetic predisposition toward lower plasma levels of two proteins - CD40 and C-C motif chemokine ligand 28 (CCL28) - was associated with increased depression risk (Figure 2A; Supplementary Table 1).

**Figure 2.**
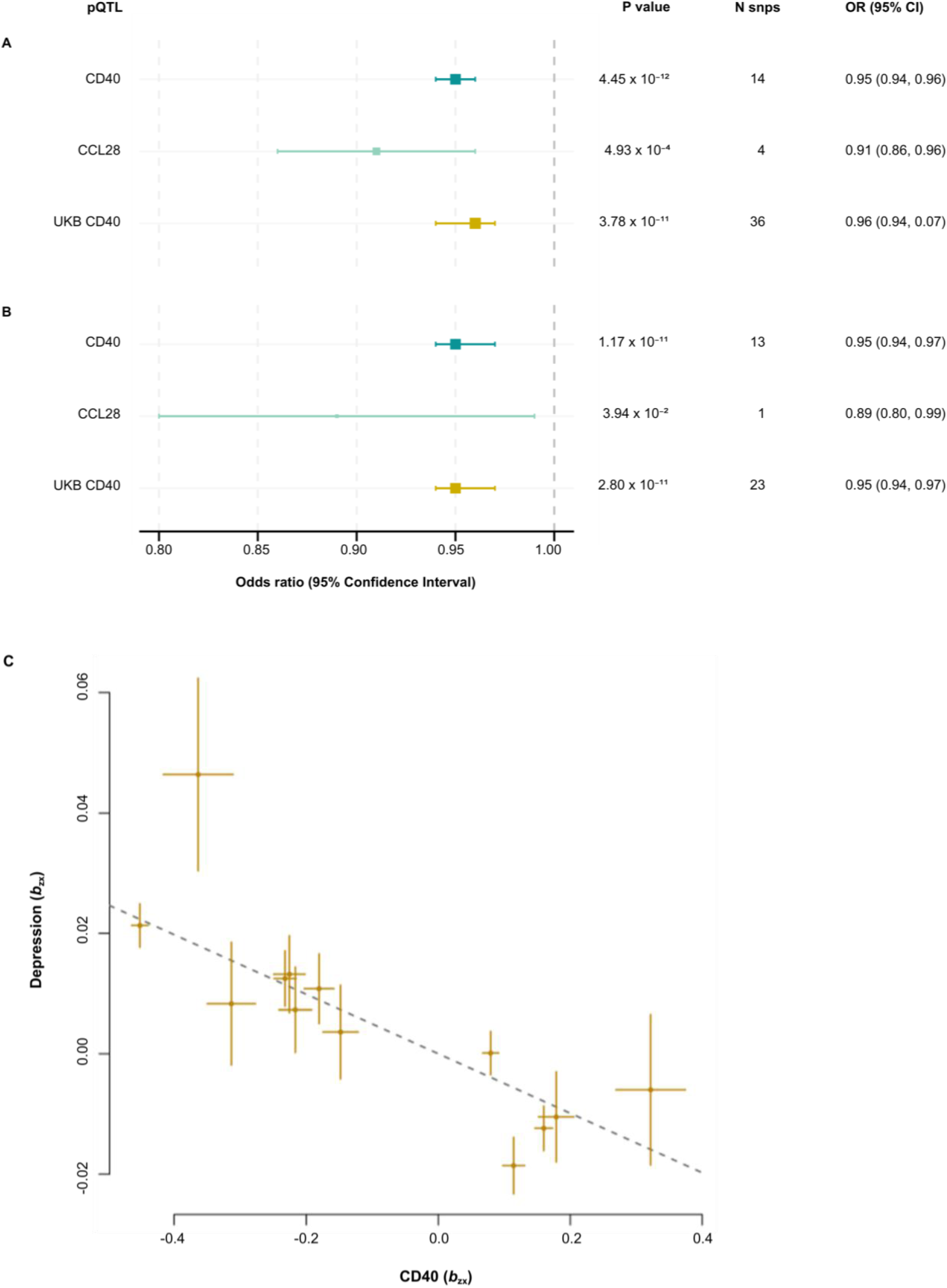
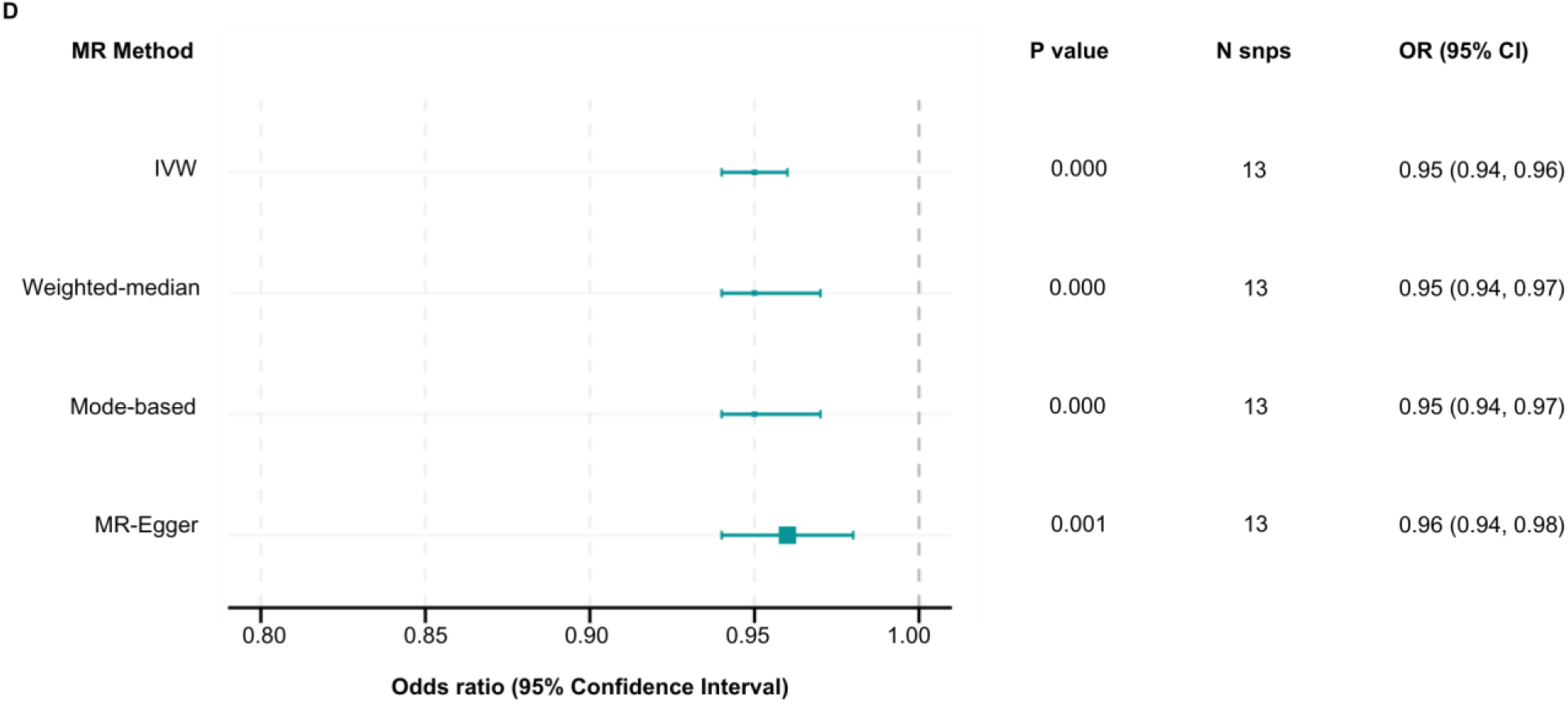
CD40 is consistently associated with depression across multiple Mendelian randomisation approaches. Bidirectional GSMR analysis identified associations between two inflammatory pQTLs and depression risk. Forest plots show odds ratio (OR) estimates for effects that passed Bonferroni correction (p_GSMR_ < 5.45 × 10⁻⁴), in both the discovery and UK Biobank (UKB) replication cohorts. Associations for the CD40 and CCL28 trans-regions were retained in both datasets **(A)**. In cis-restricted GSMR analyses designed to minimise horizontal pleiotropy **(B)**, the association with CD40 remained, while the effect for CCL28 did not reach the significance threshold. **(C)** Scatter plot of 13 independent SNP instruments in the CD40 cis-region following HEIDI-outlier filtering, with SNPs plotted by their effect on CD40 protein levels versus depression risk; error bars represent standard errors. **(D)** Forest plot summarising sensitivity analyses using IVW, MR-Egger, weighted median, and mode-based MR methods. Results support a consistent effect of CD40 on depression across all methods (*p* < 0.05). Full results in Supplementary Table 1. GSMR: generalized summary Mendelian randomisation; OR: odds ratio; pQTL: protein quantitative trait locus; UKB: UK Biobank; SNP: single nucleotide polymorphism; MR: Mendelian randomisation; IVW: inverse variance weighted.

To mitigate potential bias from horizontal pleiotropy, we repeated the analysis using only *cis*-pQTL instruments. In this sensitivity analysis, only CD40 remained significant (Figure 2B; Supplementary Table 2). Thirteen CD40 *cis*-instruments were retained after HEIDI-outlier filtering, all showing consistent direction of effect (Figure 2C; Supplementary Table 3). Reverse GSMR provided no evidence of a causal effect from depression to CD40 levels (Figure 2B; Supplementary Table 4). Additional MR sensitivity and LOO tests further confirmed the robustness of our primary findings and that no single SNP disproportionately influenced the overall signal (Figure 2D; Supplementary Table 5; Supplementary Figures 1-5). Replication using independent pQTL data from UKB supported the CD40 - depression association, with effect estimates consistent in direction and significance with those observed in the discovery cohort (Figure 2A and 2B; Supplementary Table 4).

### The association between plasma CD40 protein levels and depression is confounded by linkage disequilibrium

To determine whether plasma CD40 protein levels and depression share the same causal genetic aetiology at the *CD40* locus, we performed genetic colocalisation analyses. Pairwise Coloc analysis (see Methods) showed no robust evidence of colocalisation (PP_H4_ < 0.8), with strong support for H3 (PP_H3_ = 1.00), indicating distinct causal variants for each trait (Supplementary Table 6). The sentinel variants for CD40 protein (rs1883832) and depression (rs9074) were different, though in relatively high LD (*r*^2^ = 0.76; 1000G EUR phase 3, v5), as illustrated in the regional plot (Figure 3). Taken together, these findings suggest that the observed MR association was likely confounded by LD, and that plasma CD40 protein levels may not feature in the causal pathway for depression.

**Figure 3.**
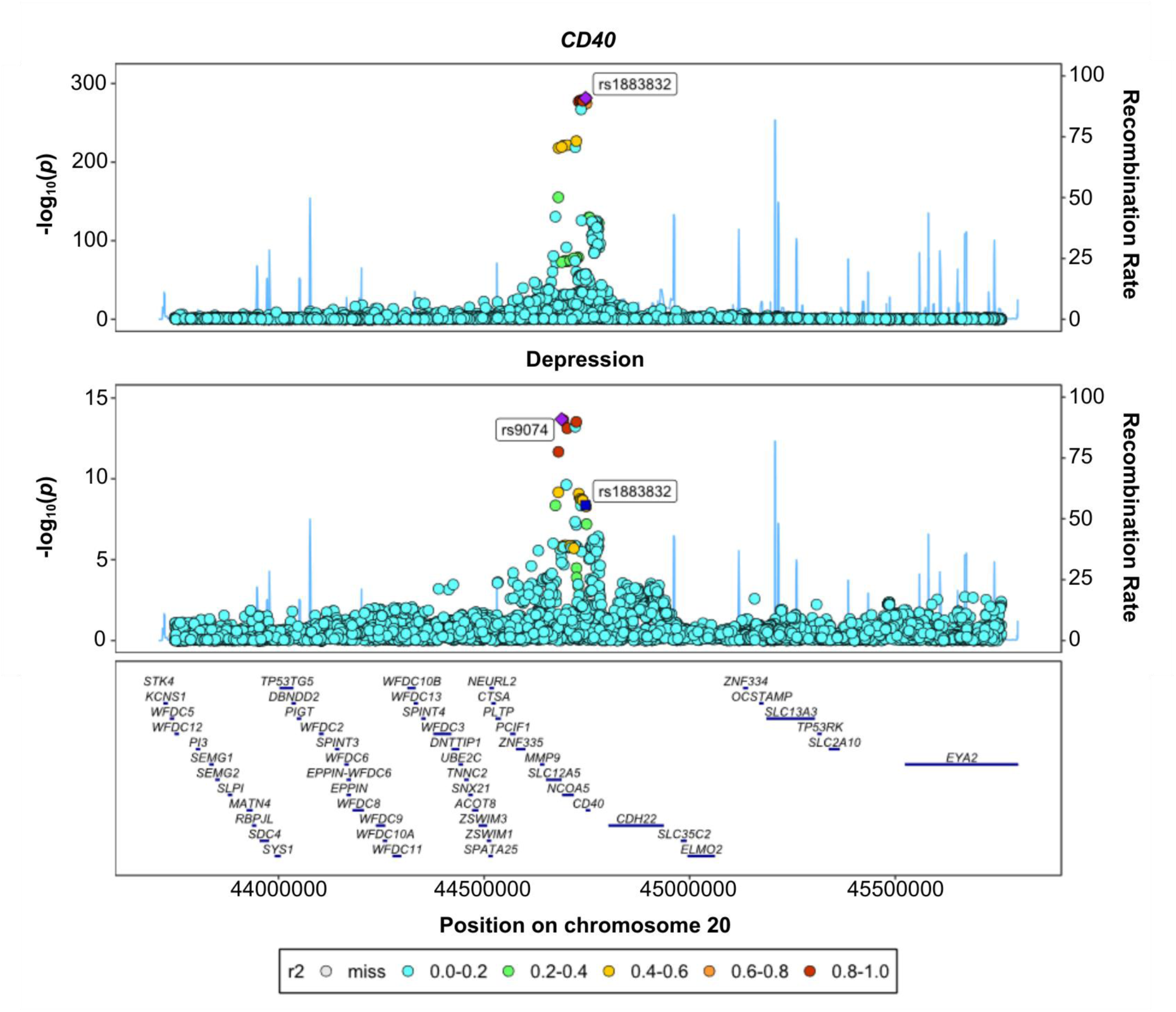
Regional association plots highlight distinct signals for CD40 protein levels and depression at the *CD40* locus. Stacked regional plots show association signals for CD40 plasma protein levels (top) and depression (bottom) across the *CD40 cis*-region. SNPs are plotted by chromosomal position (x-axis) and −log₁₀ (p-value; y-axis), with colour denoting linkage disequilibrium (LD; r²) with the lead variant. The *CD40* sentinel SNP rs1883832 is indicated in both panels.

### SLC12A5, not CD40, is the likely effector gene for depression at the locus

Given that plasma CD40 protein levels may not directly relate to depression based on these findings, we sought to identify the mechanisms driving the depression-associated signal at the *CD40* locus. We first annotated the depression sentinel variant (rs9074) and its close proxies (r^2^ > 0.8) using the Ensembl VEP (Supplementary Table 7). Next, we performed a variant look up of the depression sentinel in *cis-*eQTL data from GTEx (v8). Overall, rs9074 was associated (*p_adj_* < 0.05) with mRNA expression levels of nearby genes, two of which were protein-coding: *CD40* and solute carrier family 12 member 5 (*SLC12A5;* Supplementary Table 8). *CD40* mRNA expression was associated with rs9074 across several tissues in the brain and periphery, while *SLC12A5* mRNA was associated in brain tissue only (i.e., hippocampus, putamen, caudate). Accordingly, *SLC12A5* mRNA expression is brain-specific.

To determine which of these *cis-*eQTL signals share the same genetic aetiology with either depression or plasma CD40 protein levels at the locus, we conducted statistical colocalisation analyses. Pairwise analyses using Coloc consistently revealed a higher PP_H4_ between *CD40 cis-*eQTL signals and CD40 protein levels than for depression, whereas the opposite was true for *SLC12A5 cis-*eQTL signals (Figure 4A and 4B; Supplementary Tables 9–10).

**Figure 4:**
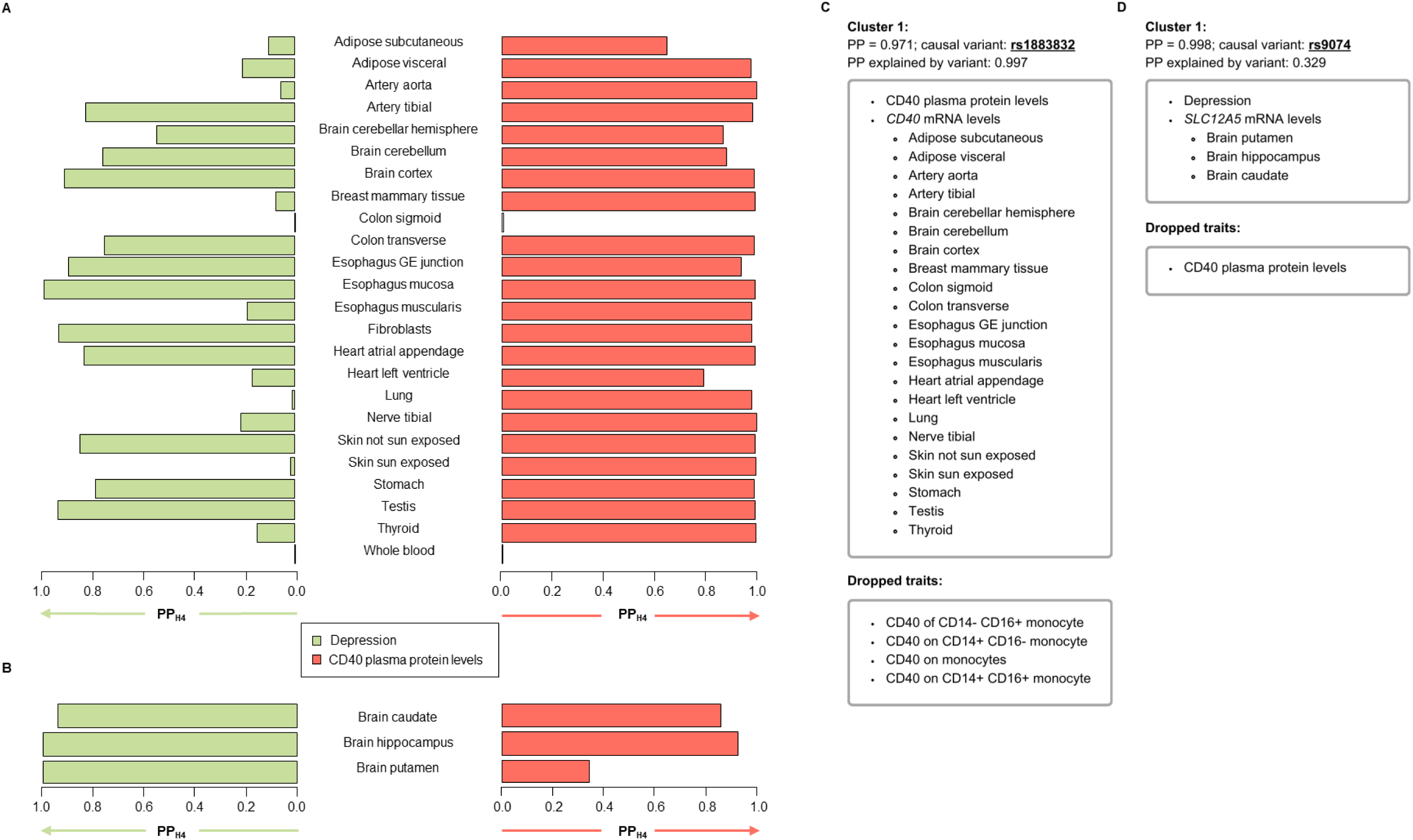
Statistical colocalisation analyses using eQTL data prioritise *SLC12A5*, not *CD40*, as the most likely effector gene at the 20q13.12 depression risk locus. Pairwise statistical colocalisation analyses (Coloc) of the depression signal (left blue bars) and the CD40 plasma protein signal (right red bars) with (A) *CD40* and (B) *SLC12A5 cis*-eQTL signals from the GTEx project (v9). *Cis*-eQTL signals for *CD40* and *SLC12A5* were selected for colocalisation analyses if they were tagged by an association with either the depression (rs9074) or CD40 plasma protein level (rs1883832) index variants according to the GTEx web portal. **(C)** Trait clusters from multi-trait statistical colocalisation analyses (Hyprcoloc) including the depression signal, the CD40 plasma protein signal, and all *CD40 cis*-eQTL signals at the 20q13.12 locus. **(D)** Trait clusters from multi-trait statistical colocalisation analyses (Hyprcoloc) including the depression signal, the CD40 plasma protein signal, and all *SLC12A5 cis*-eQTL signals at the 20q13.12 locus. The posterior probabilities (PPs) of each cluster are indicated above each cluster along with the most likely candidate causal variants and the PP they explain. PP_H4_: PP of hypothesis 4, which states that trait 1 and trait 2 share the same causal variant at a locus.

We then supplemented these Coloc analyses by applying another colocalisation method called HyPrColoc. Unlike Coloc, HyPrColoc enables multi-trait colocalisation and attempts to partition traits into clusters of colocalising traits, while any traits that fail to cluster are dropped. We first ran an iteration of HyPrColoc including depression, plasma CD40 protein levels, and all *CD40 cis-*eQTL signals. We found that CD40 protein levels clustered with the vast majority of the *CD40 cis-*eQTL signals, while depression did not cluster with any (Figure 4C; Supplementary Table 11). Conversely, in an iteration where we swapped the *CD40* for the *SLC12A5 cis-*eQTL signals, we found that depression clustered with all three *SLC12A5* signals, while the CD40 protein trait was dropped (Figure 4D; Supplementary Table 12). Taken together, these colocalisation findings suggest that the most likely effector gene for depression at the locus is *SLC12A5*, not *CD40*.

### The CD40/SLC12A5 locus modulates risk of depression and immune disease through bifurcating mechanisms

To further assess whether the CD40 and depression signals at the *CD40/SLC12A5* locus act through distinct downstream mechanisms, we conducted a PheWAS of the respective sentinel variants, followed by statistical colocalisation to evaluate shared versus distinct genetic aetiology. We identified 27 shared traits associated with both the *CD40* SNP rs1883832 and depression SNP rs9074 (Figure 5A; Supplementary Tables 13–14), as well as 5 and 6 unique traits, respectively, that reached genome-wide significance (*p* < 5 x 10^-8^). The observed overlap between many of the traits is likely driven by LD between the two sentinel variants.

**Figure 5.**
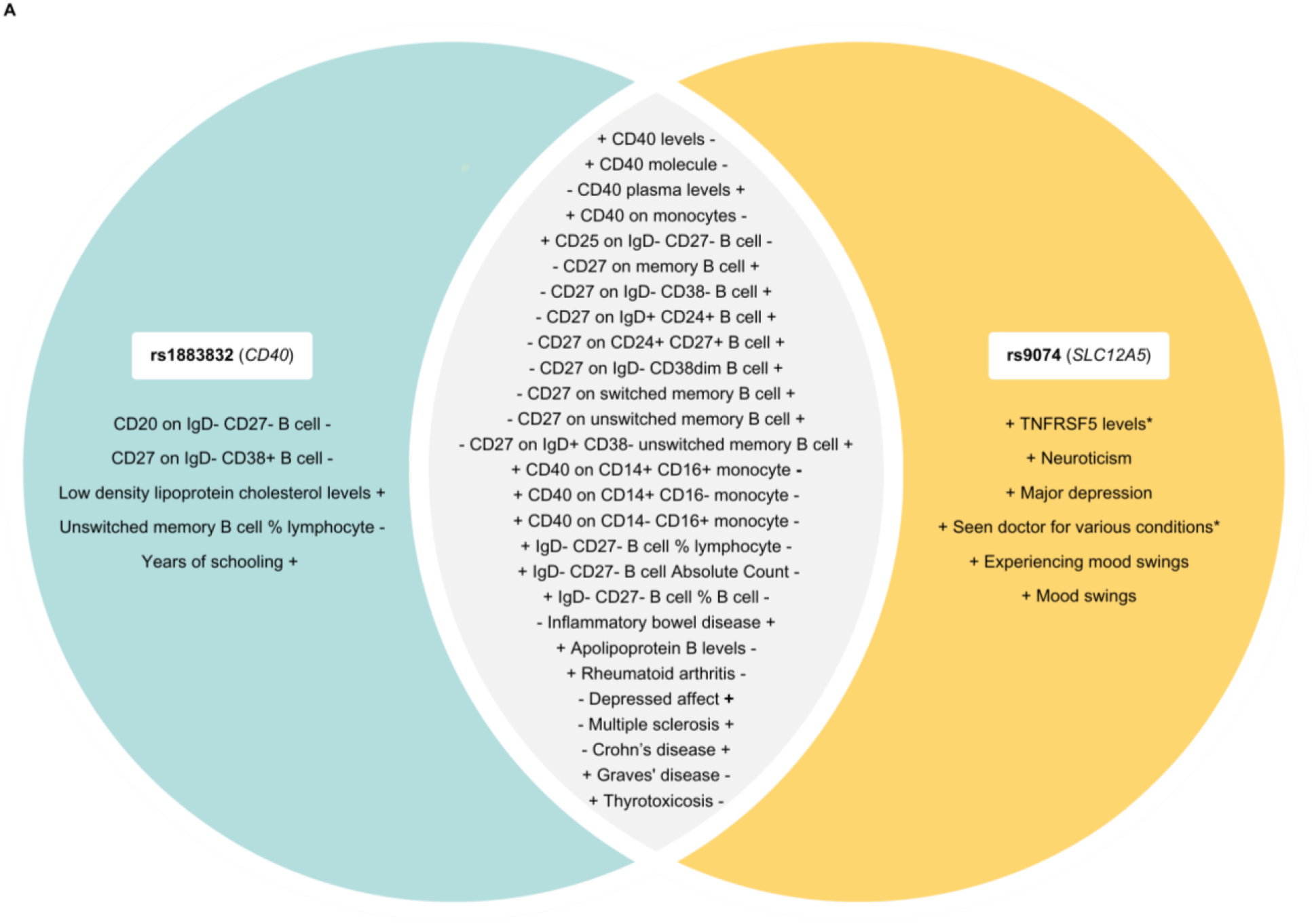
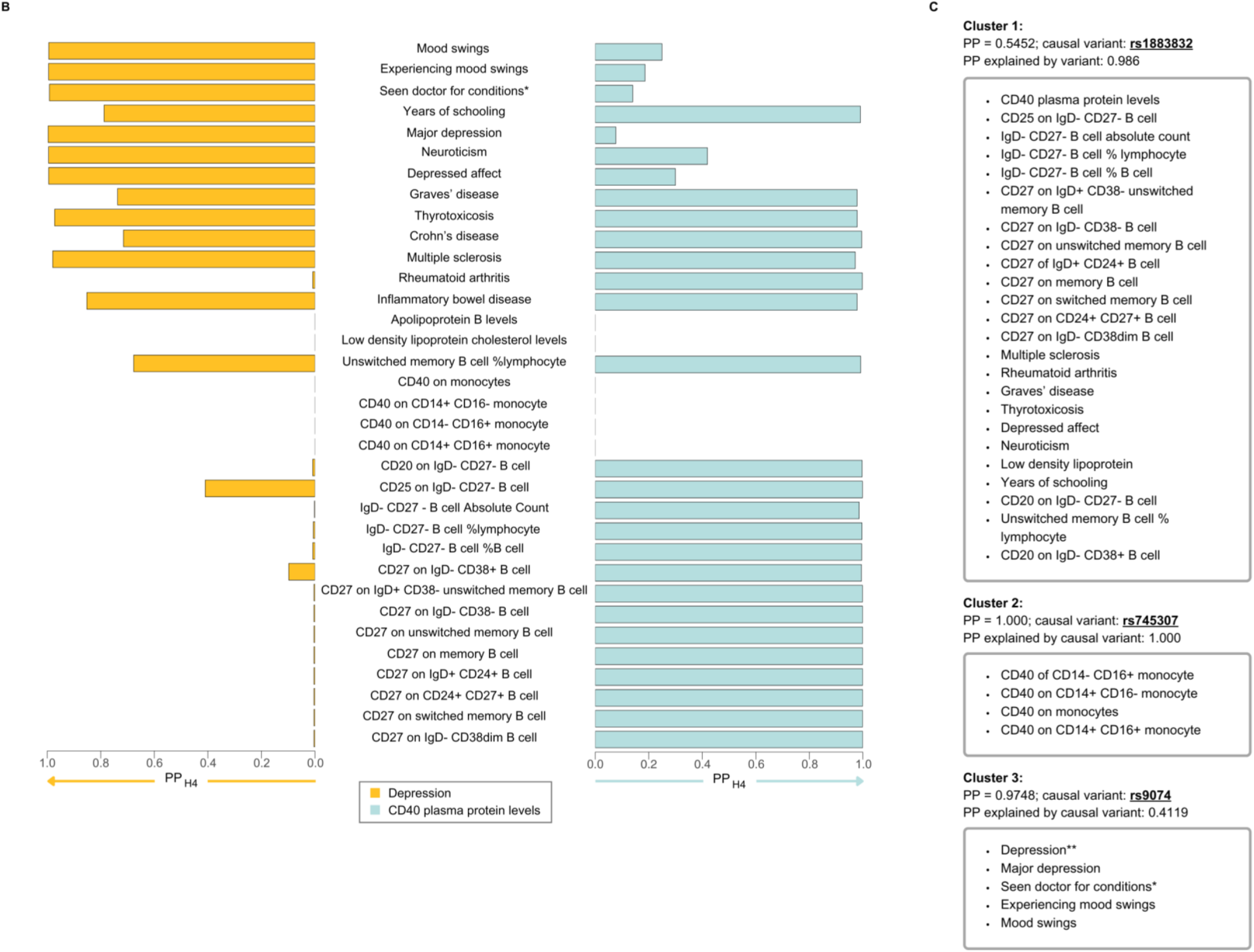
Shared and distinct trait associations at the *CD40/SLC12A5* locus implicate bifurcating immune and psychiatric pathways. **(A)** Venn diagram from PheWAS of *CD40* (rs1883832) and depression (rs9074) sentinel variants showing 27 shared, 5 CD40-specific, and 6 depression-specific genome-wide significant trait associations (p < 5 × 10⁻⁸). Trait directionalities are oriented to the CD40-elevating (rs1883832-C) and depression risk-increasing (rs9074-A) alleles. “+” and “–” denote effect direction for each SNP; ^TNFRSF5 = Tumour necrosis factor receptor superfamily member 5; *Seen doctor (GP) for nerves, anxiety, tension or depression. **(B)** Pairwise colocalisation (coloc) of each trait with CD40 protein (left) and depression (right). Immune-related traits predominantly colocalised with CD40, while psychiatric traits colocalised with depression. Several immune traits, including thyrotoxicosis, IBD, and MS, colocalised with both signals but showed opposing directional effects. **(C)** Multi-trait colocalisation (HyPrColoc) identified three clusters: one driven by CD40 plasma protein and immune traits (rs1883832), one independent monocyte-derived CD40 expression cluster (rs745307), and one psychiatric trait cluster (rs9074), with fine-mapping suggesting additional candidate variants in LD (Supplementary Table 10). **Depression: summary statistics used for discovery MR analysis.

After filtering (see Methods), we conducted pairwise colocalisation analyses between CD40-and depression-*cis* data and the corresponding traits identified in the PheWAS, together with additional relevant traits for comparison. Posterior probabilities for each trait are shown in Figure 5B and Supplementary Tables 13 –14. We found strong evidence of colocalisation (PP_H4_ > 0.8) for 9 traits with depression, and 22 traits with plasma CD40 protein levels. Traits related to immune function and disease predominantly colocalised with plasma CD40, whereas the depression signal at this locus preferentially colocalised with psychiatric traits. However, several immune-related conditions — including thyrotoxicosis, inflammatory bowel disease (IBD), and multiple sclerosis (MS) — colocalised with both signals. Notably, these traits showed opposing directional effects: the C allele for rs1883832 was associated with increased risk of thyrotoxicosis but a lower risk of inflammatory bowel disease (IBD) and multiple sclerosis (MS), whereas the A allele for rs9074 was linked to increased risk of IBD and MS but lower risk of thyrotoxicosis.

We again used HyPrColoc to disentangle shared versus distinct genetic associations across the identified traits. HyPrColoc revealed three distinct clusters of colocalised traits (Figure 5C; Supplementary Table 15). The first cluster comprised 24 traits, including CD40 plasma protein levels, immune-related diseases, and cellular traits, and showed moderate colocalisation with rs1883832. The second cluster showed robust colocalisation with rs745307, which fully explained the posterior probability. This signal appeared independent of both rs1883832 and rs9074, suggesting that the genetic signal for CD40 expression in monocyte subsets — including classical (CD14+CD16-), non-classical (CD14-CD16+), and intermediate (CD14+CD16+) monocytes — is distinct from depression and other immune-related traits. These findings imply that myeloid-derived CD40 expression may be less relevant to immune-mediated disease compared to lymphoid-derived signals. The third cluster, comprising depression and mood traits, colocalised with sentinel variant rs9074, although this variant accounted for only 41.2% of the posterior probability. Fine-mapping within this cluster (Supplementary Table 15) suggested that rs13037326 (PP_H4_ = 0.3761) and rs6131010 (PP_H4_ = 0.1760), which are in perfect LD with rs9074 (*r*^2^ = 1), may also contribute to the association, but with weaker evidence compared to rs9074.

## Discussion

We applied GSMR to systematically analyse 91 immune-related pQTLs for potential causal roles in depression. We identified an association between CD40 and depression, which was robust across multiple testing and sensitivity analyses. However, colocalisation was not observed, suggesting that the link may be due to pleiotropic effects at the *CD40* locus, with *SLC12A5* likely acting as the effector gene for depression (see Figure 6). Further functional and PheWAS analyses supported this interpretation, showing that the CD40 protein signal (rs1883832) was primarily associated with inflammatory disorders, while the depression signal at the *CD40/SLC12A5* locus (rs9074) showed stronger ties to psychiatric traits. Additionally, we found no evidence for reverse causality between depression and CD40 protein levels, indicating that reverse causality is unlikely to explain the inflammation observed in this population.

**Figure 6.**
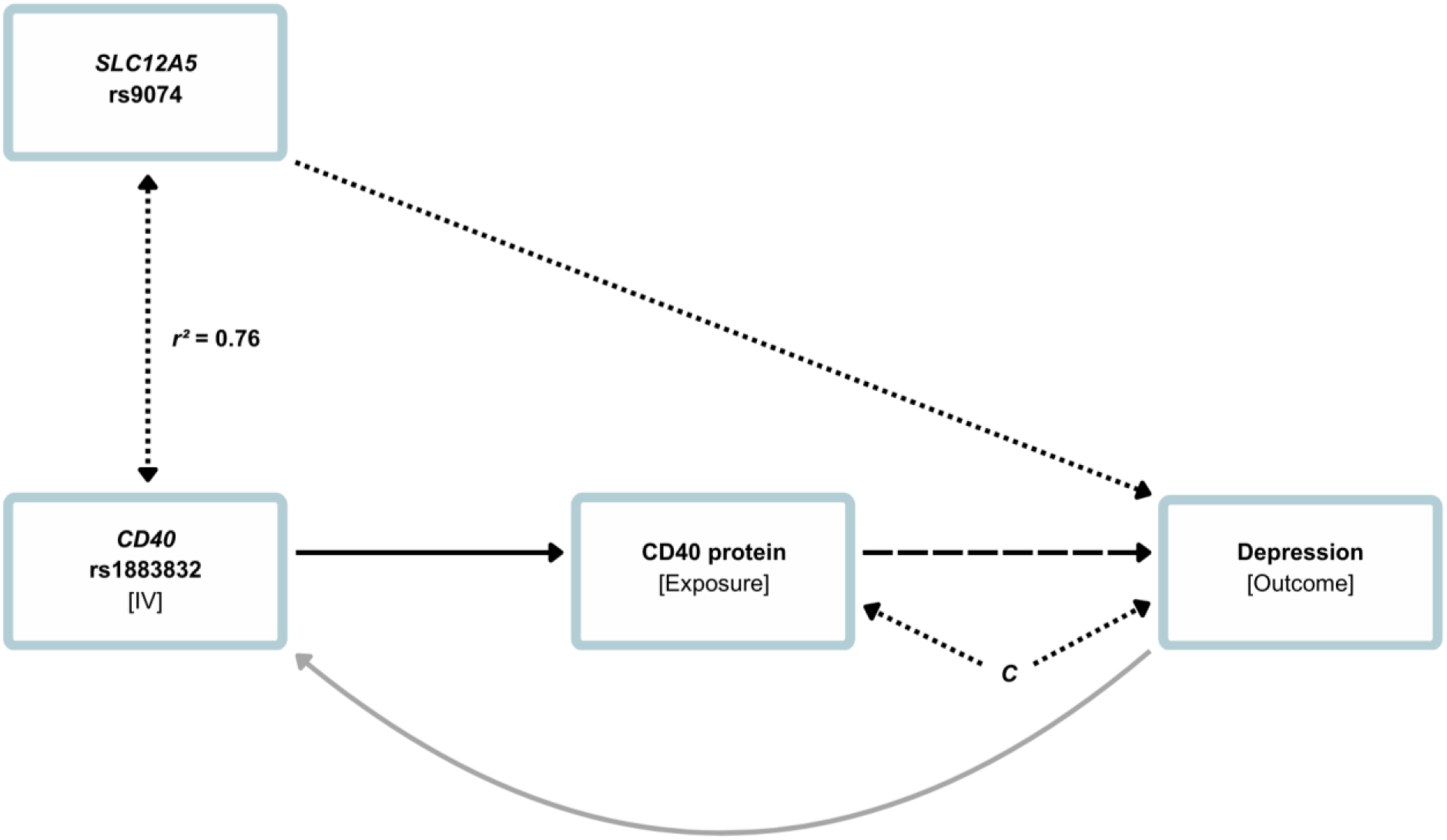
Pathway diagram illustrating the Mendelian randomisation (MR) framework used to investigate the potential causal relationship between CD40 protein levels and depression. The *CD40* sentinel variant rs9074 serves as the primary instrumental variable (IV) for CD40 protein levels, while *SLC12A5* variant rs9074 represents a possible pleiotropic association with the IV, given its high linkage disequilibrium (LD) with rs1883832 and its functional relevance to depression. The solid black arrow shows the assumed causal effect of the IV on the exposure, while the dashed arrow denotes the hypothesised causal relationship between CD40 protein and depression, which was not supported by colocalisation analysis. Dotted arrows indicate potential associations, including between *SLC12A5* and depression, as well as between *SLC12A5* and *CD40* via LD. The grey arrow illustrates the lack of evidence for a reverse causal effect of depression on CD40 levels. IV: instrumental variable; *C*: unmeasured confounders.

Evidence from well-powered GWAS indicate that pleiotropy across complex traits is extensive, with up to 90% of observed effects arising from locus-level pleiotropy [47]. This can occur either when a single gene influences multiple traits, or when distinct genes or SNPs within the same locus affect different traits due to LD, implicating the same region overall. Such pleiotropy can inflate causal estimates and lead to type I errors in MR, violating the exchangeability assumption [48]. While LD confounding is recognised, it is often overlooked in practice [49] and can go undetected when using conventional MR methods. Colocalisation provides a solution to this issue as it generally requires stronger evidence to support causal assumptions than MR [48] and represents a strength of this study. Indeed, a recent phenome-wide MR study examining plasma pQTLs (including *cis* and *trans* instruments) across various complex disorders found that over 30% of associations identified through MR did not colocalise, indicating that genetic confounding due to LD is likely widespread in pQTL MR studies [40].

The conflicting results from MR and colocalisation in our study suggest potential genetic confounding from LD between SNPs at the *CD40* locus. Here, rs1883832, initially flagged by MR as a causal variant, is in high LD (r² = 0.76) with rs9074, a variant located within the *SLC12A5* gene region. This suggests that the association with depression may be mediated via regulatory effects on *SLC12A5*, rather than *CD40*. This highlights the challenge posed by locus pleiotropy in the context of MR, where distinct causal variants at the same locus influence different traits through shared LD mechanisms [50]. This is complicated even further at the *CD40*/*SLC12A5* locus due to the moderate to high LD observed between the CD40 and depression sentinel variants at the locus. Similar patterns of distinct causal variants within both genes and loci have been observed in MR analyses of other traits and disorders [48].

The LD between depression-linked and *CD40* variants demonstrates an association that could influence immune profiles in ways that may be clinically relevant, but unlikely to be causally relevant for depression. This is especially applicable to personalised care in depression, given that pleiotropic effects within other immune loci (e.g., the HLA region) are implicated in various neuropsychiatric and immune-related outcomes [51, 52]. *CD40* itself is a stimulatory receptor linked to immune cell proliferation and cytokine production [53, 54] and is associated with a broad spectrum of inflammatory, autoimmune [55–58], and cognitive traits, including educational attainment and Alzheimer’s disease [59, 60]. Some evidence indicates that these associations may arise from both specific and broad pleiotropic effects, as variants associated with *CD40* are frequently in high LD or shared across multiple traits [61–63]. The effects of *CD40* on various phenotypes also appear heterogeneous, with plasma levels associated with an increased risk for some immune disorders and decreased risk for others [30].

Prior MR studies investigating inflammatory links to depression have yielded mixed findings. While some support causal associations with immune-related proteins such as CRP and IL-6 (e.g., [64]), others do not (e.g., [28, 65]). Such inconsistencies often stem from differences in statistical power, methodology, and instrumental variable selection (e.g., GWAS-derived pQTLs versus candidate SNPs) [66]. Therefore, triangulating complementary approaches in MR studies (e.g., MR, colocalisation, functional analyses) is essential to minimising bias and strengthening causal inferences [67].

While no study has established a causal relationship between *CD40* and depression, research implicates dysregulation of CD40 protein and its ligand (CD40L) in major depressive disorder (MDD) and other neurological conditions [68–74]. Notably, our initial findings indicate that lower genetically predicted CD40 levels are associated with increased depression risk. This contrasts with several observational studies showing elevated CD40 levels in depressed individuals [68–71] but aligns with one other report [72]. A similar discrepancy also appears between CRP levels and schizophrenia, where inverse genetic associations challenge conventional inflammatory hypotheses [75]. Such findings raise the possibility that low-grade inflammation, including altered CD40 levels, may reflect downstream consequences of psychiatric pathophysiology rather than primary causal mechanisms. Furthermore, the functional polymorphism, rs1883832, which is associated with several inflammatory disorders such as coronary heart disease [76] – a condition frequently comorbid with depression [64, 77–79] – may instead represent a plausible mechanistic link between depression and immune comorbidities.

Our GTEx analyses indicated *SLC12A5* as the likely effector gene for depression, following the colocalisation of *cis*-eQTL signals with this trait. *SLC12A5*, also known as *KCC2*, encodes a potassium-chloride co-transporter that regulates intracellular chloride levels and is primarily expressed in neurons. Given its role in maintaining the chloride gradient necessary for hyperpolarizing inhibition mediated by γ-aminobutyric acid (GABA) and its links to neurodevelopmental conditions [80, 81], genetic variation at this locus may influence depression risk via altered inhibitory tone and downstream effects on hypothalamic-pituitary-adrenal (HPA) axis function [82, 83]. *SLC12A5* expression has been associated with psychiatric traits including depression, neuroticism, schizophrenia, and autism [84–87] and is linked to brain structure and function [88]. It is also implicated in neurological conditions, including paediatric epilepsy and multiple sclerosis (MS) [89, 90]. Colocalisation analyses in MS have identified *SLC12A5* expression in both excitatory and inhibitory neurons at the *CD40* locus-one of only 11 loci exclusively colocalised in brain cell types rather than immune cells - suggesting a neuron-specific role for this locus in some traits [91]. The *CD40* locus also functions as a *cis*-eQTL for B cells and is associated with rheumatoid arthritis susceptibility [92, 93], indicating that distinct mechanisms may operate in different cell types. The potential relevance of this locus in depression is further supported by a recent GWAS of anxiety, which implicated both *CD40* and *SLC12A5* as potential effector genes and highlighted the importance of GABAergic signalling, which is particularly salient given the substantial genetic correlation between anxiety and depression [94]. Together with results from the present study, these findings reinforce *CD40*/*SLC12A5* as an important pleiotropic locus with distinct roles across immune and psychiatric traits.

Although this study has several strengths, certain limitations should be acknowledged. Phenotypic heterogeneity poses a significant challenge, particularly for polygenic disorders like depression, which encompass a diverse range of symptoms that vary across individuals. This variability may weaken associations in heterogeneous GWAS samples, as inflammatory risk factors could be more relevant to specific depressive subtypes [95, 96]. Additionally, depression linked to chronic or acute illnesses or neurodevelopmental conditions associated with *SLC12A5* (e.g., epilepsy, cancer, neuropathic pain) may further confound genetic associations. The reliance on binary phenotypes, such as “depression,” also presents challenges for causal inference. Dichotomizing a continuous trait like depression symptoms may not fully capture the complexity of the disorder, which can introduce biases and complicate clinical interpretations. A critical issue is also the focus on individuals of predominantly Caucasian ancestry with self-reported depression limiting generalizability. Further studies are needed to explore the relevance of these findings across a broader range of ancestries, as well as different diagnostic criteria and disease states (e.g., depression progression versus incidence).

In conclusion, MR analyses indicated a potential association between plasma CD40 protein levels and depression, however colocalisation analyses indicated this finding was confounded by LD. These findings are therefore not consistent with a causal role for *CD40* in depression. Instead, we demonstrated for the first time a pleiotropic effect mediated by *SLC12A5*. This warrants further investigation in independent datasets and cohorts where functional analyses may be conducted. Our results illustrate the importance of combining MR and colocalisation analyses to clarify associations in traits with complex genetic architecture, such as depression, where pleiotropic effects and multiple pathways may be involved. Further functional and multi-omic research is essential to further define the roles of *SLC12A5* and *CD40* in depression pathophysiology and immune comorbidities, respectively.

## Data Availability

All data produced in this work (summary level MR, colocalisation, and HyPrColoc outputs) are provided in the manuscript and Supplementary Material. Source summary statistics analysed were obtained from publicly available repositories cited in the manuscript; no new human data were collected.

https://ipsych.dk/en/research/downloads/

https://www.phpc.cam.ac.uk/ceu/proteins

http://ukb-ppp.gwas.eu

https://www.gtexportal.org

http://gwas-api.mrcieu.ac.uk

https://www.internationalgenome.org/

## Acknowledgments

E.H. is supported by the National Health and Medical Research Council (NHMRC) Leadership Investigator Award (GNT2025349). We thank the investigators and participants of the contributing GWAS and QTL studies for making the data publicly available.

