## Supplementary Figures 1 - 5 for "Mendelian randomisation and colocalisation reveal pleiotropic effects of *CD40/SLC12A5* locus on CD40 protein, depression, and immune disease"

**Contents**

This file contains Supplementary Figures 1–5 showing scatter plots, variant-level estimates, and leave-one-out results for the CD40–depression sensitivity analyses.

1. **Supplementary Figure 1.** Scatter plot of genetic associations for CD40 pQTL instruments versus depression; IVW regression line shown.
2. **Supplementary Figure 2.** Sensitivity plots across multiple Mendelian randomization methods (median, IVW, penalized/robust variants, MR-Egger), demonstrating consistency of the CD40–depression association.
3. **Supplementary Figure 3.** Variant-level IVW causal estimates (95% CI) with pooled IVW estimate.
4. **Supplementary Figure 4.** Variant-level MR-Egger causal estimates (95% CI) with pooled MR-Egger estimate.
5. **Supplementary Figure 5.** Leave-one-out IVW analysis (99% CI) indicating no single SNP drives the overall CD40–depression effect.

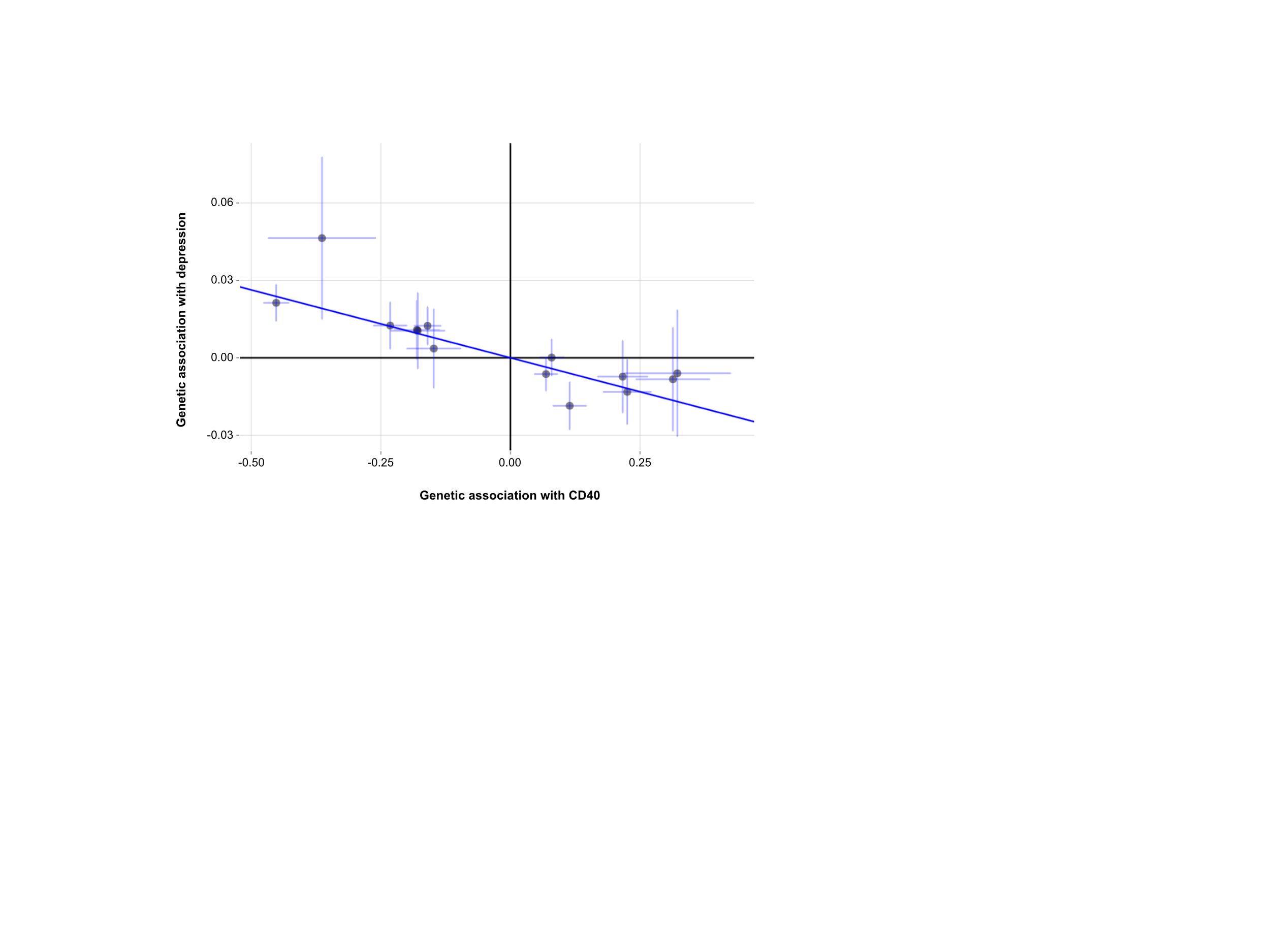

**Supplementary Figure 1. Scatter plot of genetic association between CD40 and depression.** The plot displays the genetic associations of 13 independent CD40 pQTL instruments (x-axis) against their corresponding associations with depression risk (y-axis). Each point represents a single genetic variant, with horizontal and vertical lines indicating standard errors. The blue regression line represents the inverse variance-weighted (IVW) Mendelian randomisation estimate. The observed negative slope suggests a potential causal relationship between CD40 protein levels and reduced MDD risk.

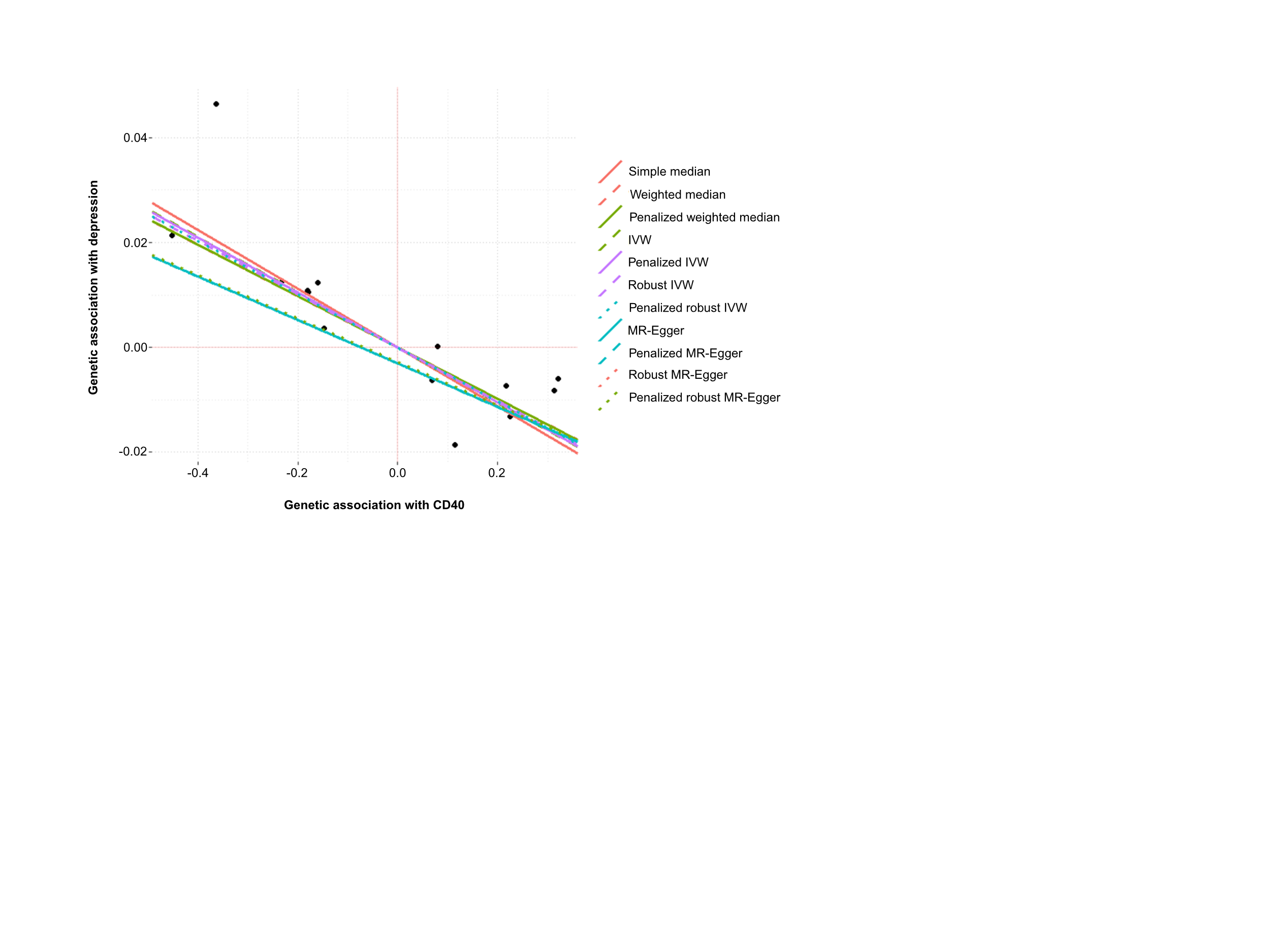

**Supplementary Figure 2**. **Sensitivity analyses of genetic associations between CD40 and MDD using multiple Mendelian randomization methods.** The plot demonstrates the consistency of the negative association between CD40 and depression across different methodological approaches. Methods included simple median, weighted median, penalized weighted median, inverse variance weighted (IVW), penalized IVW, robust IVW, penalized robust IVW, MR-Egger, penalized MR-Egger, robust MR-Egger, and penalized robust MR-Egger regression. Black dots represent individual CD40 instruments. The x-axis shows the genetic association with CD40 protein levels, while the y-axis shows the genetic association with risk for MDD. The convergence of results across multiple methods supports the primary finding and minimal impact of horizontal pleiotropy.

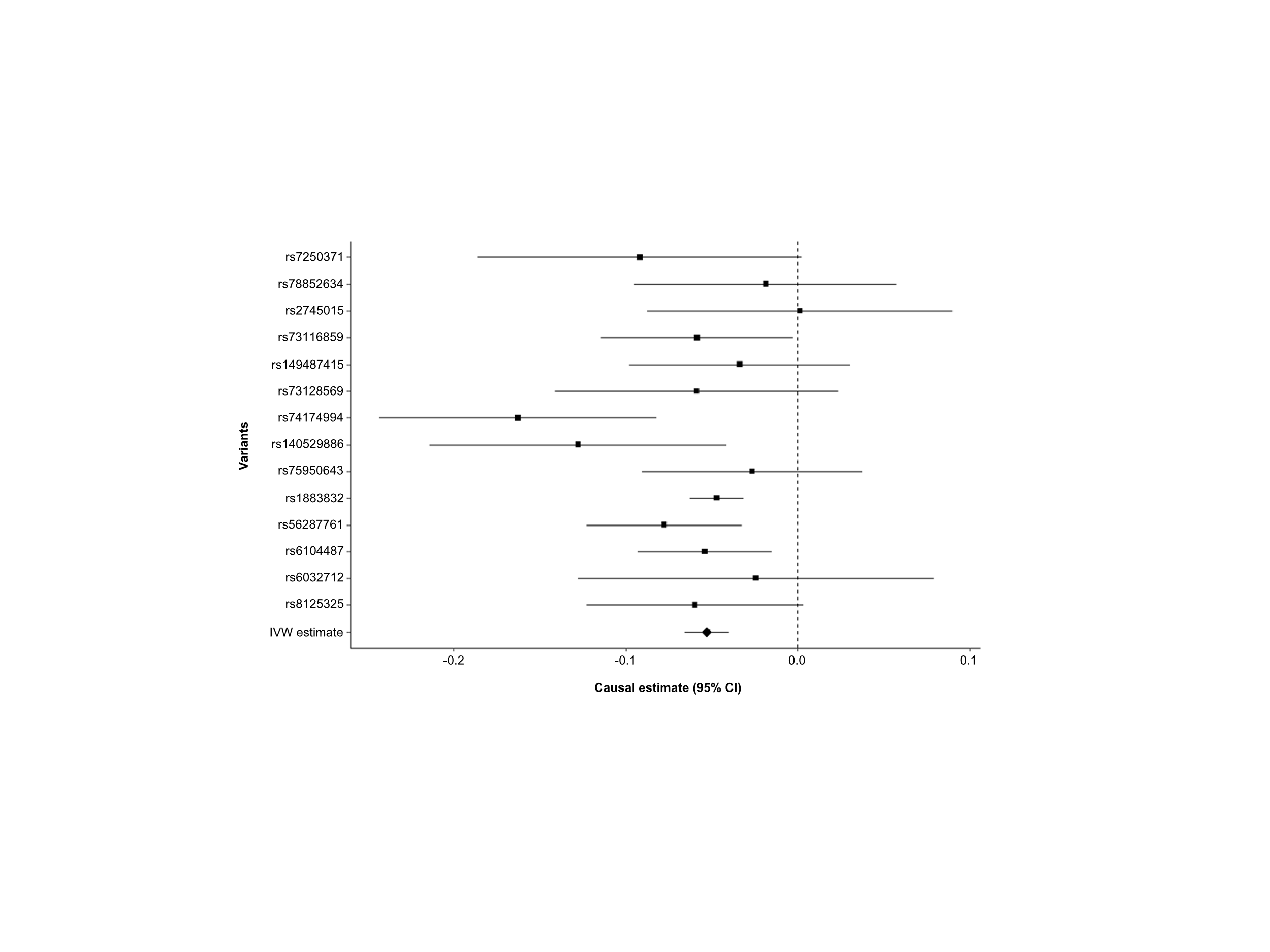

**Supplementary Figure 3. Inverse variance-weighted (IVW) causal estimates for the effect of *CD40* variants on depression risk.** Each black square represents the IVW causal estimate for an individual *CD40* variant, with horizontal lines indicating the 95% confidence interval. The diamond at the bottom represents the overall IVW estimate using all variants.

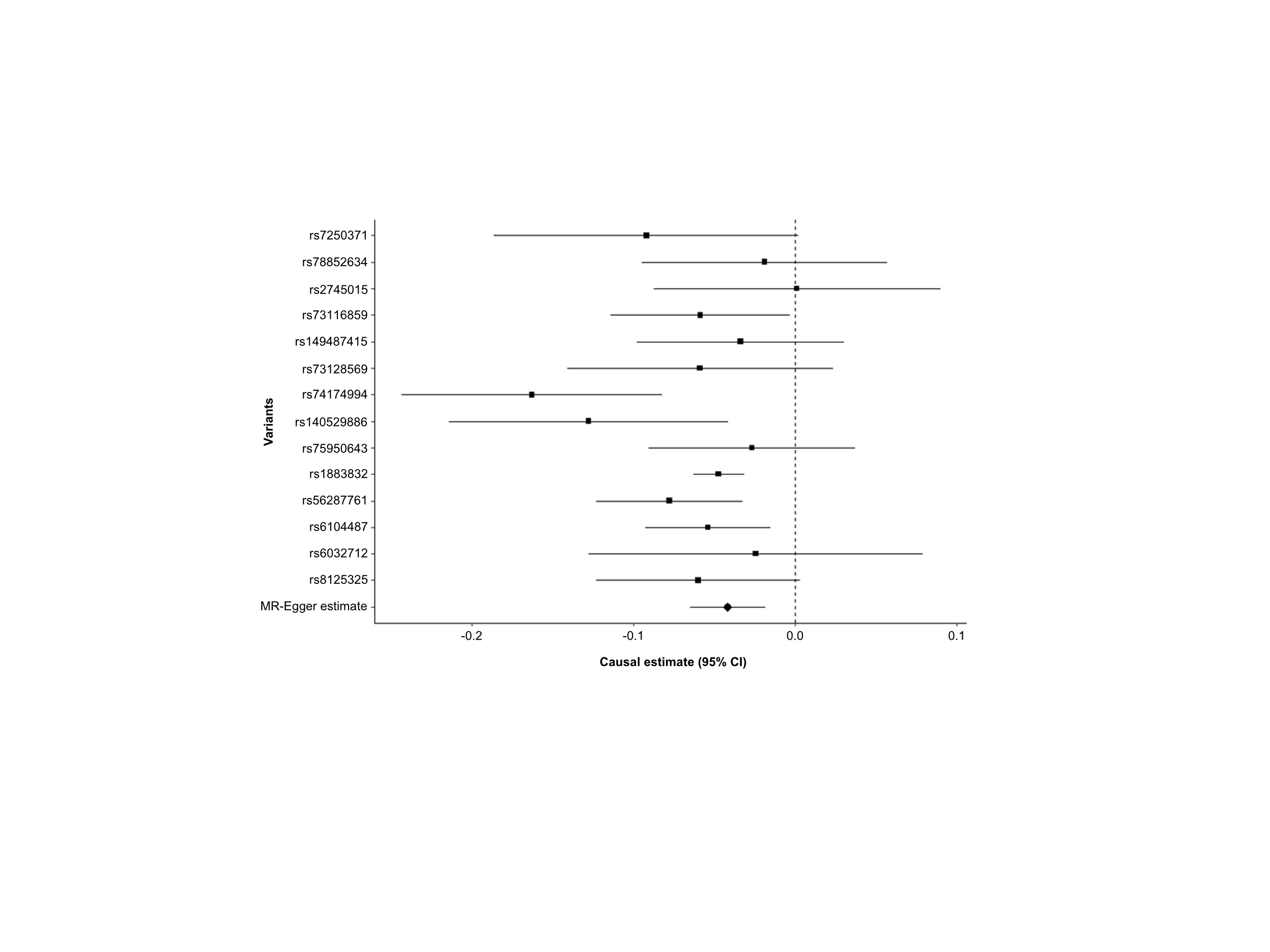

**Supplementary Figure 4.** **MR-Egger causal estimates for the effect of *CD40* variants on depression risk.** Each black square represents the MR-Egger causal estimate for an individual *CD40* variant, with horizontal lines indicating the 95% confidence interval. The diamond at the bottom represents the overall MR-Egger estimate using all variants.

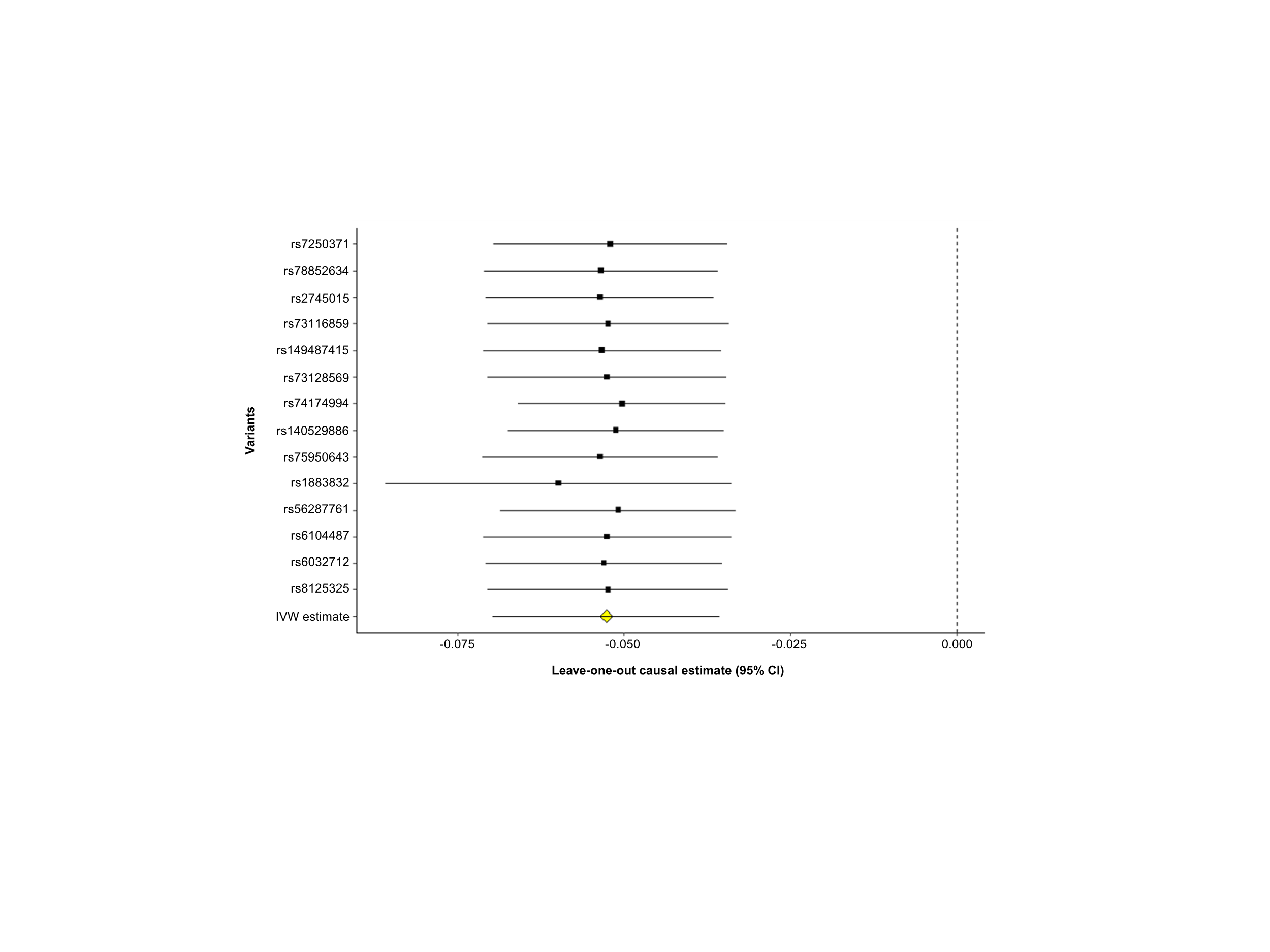

**Supplementary Figure 5. Leave-one-out (LOO) sensitivity analysis for the causal effect of *CD40* on depression.** Each black square represents the inverse variance-weighted (IVW) causal estimate obtained after omitting a single variant (labelled), with horizontal lines indicating the 99% confidence interval. The yellow diamond represents the overall IVW estimate using all variants. Combined estimates indicate that no single SNP disproportionately influenced the overall causal effect.
